# The genetic and clinical landscape of nanophthalmos in an Australian cohort

**DOI:** 10.1101/19013599

**Authors:** Owen M Siggs, Mona S Awadalla, Emmanuelle Souzeau, Sandra E Staffieri, Lisa S Kearns, Kate Laurie, Abraham Kuot, Ayub Qassim, Thomas L Edwards, Michael A Coote, Erica Mancel, Mark J Walland, Joanne Dondey, Anna Galanopoulous, Robert J Casson, Richard A Mills, Daniel G MacArthur, Jonathan B Ruddle, Kathryn P Burdon, Jamie E Craig

**Affiliations:** Department of Ophthalmology, Flinders University, Adelaide, Australia; Centre for Eye Research Australia, Royal Victorian Eye and Ear Hospital, Melbourne, Australia; Department of Ophthalmology, University of Melbourne, Melbourne, Australia; Department of Ophthalmology, Royal Children’s Hospital, Melbourne, Australia; Centre Hospitalier Territorial de Nouvelle-Calédonie, Noumea, New Caledonia; Glaucoma Investigation and Research Unit, Royal Victorian Eye and Ear Hospital, Melbourne, Australia; Royal Victorian Eye and Ear Hospital, Melbourne, Australia; Discipline of Ophthalmology & Visual Sciences, University of Adelaide, Adelaide, Australia; Program in Medical and Population Genetics, Broad Institute of MIT and Harvard, Boston, Massachusetts, USA; Analytic and Translational Genetics Unit, Massachusetts General Hospital, Boston, Massachusetts, USA; Menzies Institute for Medical Research, University of Tasmania, Hobart, Australia

**Keywords:** microphthalmia, nanophthalmos, posterior microphthalmos, axial length, *PRSS56*, *MFRP*, *TMEM98*, *MYRF*

## Abstract

**Purpose:** Refractive error is caused by a disparity between the axial length and focusing power of the eye. Nanophthalmos is a rare ocular abnormality in which both eyes are abnormally small, typically causing extreme hyperopic refractive error, and associated with an increased risk of angle-closure glaucoma.

**Methods:** A cohort of 40 individuals from 13 unrelated nanophthalmos kindreds were recruited, with 11 probands subjected to exome sequencing.

**Results:** Nine probands (69.2%) were assigned a genetic diagnosis, with variants in *PRSS56* (4), *MFRP* (3), and previously reported variants in *TMEM98* (1) and *MYRF* (1). Two of the four *PRSS56* probands harboured the previously described c.1066dupC frameshift variant implicated in over half of all reported *PRSS56* kindreds, with surrounding haplotypes distinct from each other, and from a previously reported Tunisian c.1066dupC haplotype. Individuals with a genetic diagnosis had shorter mean axial lengths (*P*=7.22×10^−9^) and more extreme hyperopia (*P*=0.0005) than those without a genetic diagnosis, with recessive forms associated with the shortest axial lengths and highest hyperopia. All individuals with an axial length below 18 mm in their smaller eye (17/17) were assigned a genetic diagnosis.

**Conclusions:** These findings detail the genetic architecture of nanophthalmos in an Australian cohort of predominantly European ancestry, their relative clinical phenotypes, and highlight the shared genetic architecture of rare and common disorders of refractive error.

## Introduction

Clear vision requires precise regulation of ocular growth, such that the axial length of the eye matches the optical focal plane created by the cornea and lens. A discrepancy between the two results in refractive error, which is the leading cause of visual impairment and the second leading cause of blindness worldwide.^1^ Refractive error may be termed either myopia (where the focal point is in front of the retina) or hyperopia (where it falls behind it). Both forms of refractive error are correctable, although individuals at the extremes of refractive error are at an increased risk of irreversible blindness due to associated complications such as myopic macular degeneration, retinal detachment, or primary open-angle glaucoma (in the case of myopia); or strabismus, amblyopia, retinal detachment and angle-closure glaucoma (in the case of hyperopia).^2^

Important insight into the developmental regulation of ocular axial length has come from the study of microphthalmia, which affects approximately 1 in 10,000 live births.^3^ Nanophthalmos and posterior microphthalmos are two rare subtypes of microphthalmia, both of which are associated with reduced axial length and high hyperopia.^4^ Posterior microphthalmos is characterised by reduced length of the posterior segment of the eye, with the anterior segment typically of normal length. Patients with posterior microphthalmos usually present with reduced visual acuity from an increase in the papillomacular retinal fold, occasional pigmentary retinopathy, and are susceptible to uveal effusions.^5,6^ Unlike posterior microphthalmos, nanophthalmos is characterized by a decrease in the size of both posterior and anterior segments, with a predisposition to primary angle-closure glaucoma.^5,6^ However, both posterior microphthalmos and nanophthalmos can be allelic^7^, supporting the hypothesis that they represent a continuum of the same phenotypic spectrum, rather than two distinct entities.^8^

Posterior microphthalmos and nanophthalmos are commonly inherited as autosomal recessive traits, with variants in two genes accounting for the majority of known cases. The first of these, *MFRP* (membrane frizzled-related protein), is expressed in retinal pigment epithelium and ciliary body, where a lack of MFRP disrupts emmetropization through an uncertain mechanism.^9,10^ Rare biallelic *MFRP* variants have been associated with nanophthalmos^11^, isolated posterior microphthalmos^7^, and posterior microphthalmos with retinitis pigmentosa, foveoschisis, and optic disc drusen^12^. Both MFRP-deficient mice^13^ and zebrafish^14^ have hyperopic phenotypes, consistent with a presumed loss-of-function mechanism in humans^11^. Viral transduction of wild-type *Mfrp* into 5 day-old *Mfrp* mutant mice can also restore axial length^13^, suggesting that MFRP provides support for ocular development both *in utero* and after birth.

A second gene, *PRSS56* (protease, serine, 56), was initially identified in autosomal recessive nanophthalmos and posterior microphthalmos kindreds from the Faroe Islands, Tunisia, Canada, and Mexico.^15–17^ A chemically-induced mouse mutant at the orthologous locus showed close phenotypic parallels, with shortened axial length, hyperopia, and a propensity for angle closure.^17^ Recent work has shown that *PRSS56* is derived from late retinal progenitor cells or Müller glia, and promotes elongation of ocular axial length both before and after eye opening.^18^ *PRSS56* variants have also been implicated in multiple independent genome-wide association studies of myopia.^19–21^

Autosomal dominant nanophthalmos is a less common clinical entity, and until recently only associated with variants in *TMEM98*^*22,23*^. Like *PRSS56, TMEM98* has also been associated with myopia in a large genome-wide association study.^21^ A second dominant nanophthalmos locus, previously mapped to chromosome 11^24^, has now been associated with variants in *MYRF*, which encodes a transcriptional regulator that drives expression of *TMEM98*.^25–28^

Here we assembled a cohort of 40 individuals across 13 nanophthalmos kindreds, describe their underlying genetic cause, and detail the relative clinical presentations associated with each gene.

## Methods

### Subjects

Individuals with nanophthalmos were recruited through the Australian and New Zealand Registry of Advanced Glaucoma^29^, and were included if they presented with bilateral and symmetrically small eyes, with an axial length less than 20 mm in both eyes. This threshold of 20 mm was previously shown to account for individuals below 3 standard deviations from the mean in the EPIC-Norfolk Eye Study, and proposed as a definition for nanophthalmos in the absence of any other standardised definition.^30^ Exclusion criteria included unilateral disease, anterior segment coloboma, and multiple non-ophthalmic syndromic features. Angle closure glaucoma status was assessed in all participants, yet was not a criterion for inclusion. Ocular hypertension was defined as an intraocular pressure (IOP) above 21 mmHg, with glaucoma defined as a glaucomatous visual field defect with a corresponding neuroretinal rim thinning. Previously reported families with variants in *TMEM98* and *MYRF* were included in this cohort.^23,28^ Research followed the tenets of the Declaration of Helsinki with informed consent obtained from the subjects after explanation of the nature and possible consequences of the study. Ethics approval was granted by the Southern Adelaide Clinical Human Research Ethics Committee.

### Sequencing

Exome sequencing and analysis was performed as previously described.^31^ Briefly, genomic DNA was extracted from venous blood samples using a QiaAmp DNA blood Maxi Kit (Qiagen), or from saliva using Oragene saliva DNA collection kits (DNA Genotek) according to the manufacturers’ protocols. Exome capture was performed using the Agilent SureSelect system (v4 or v5) and paired-end libraries sequenced on an Illumina HiSeq (2000, 2500, or 4000). Read alignment and variant calling were performed according to GATK best practices for germline short variant discovery (https://software.broadinstitute.org/gatk/). Variant annotation was performed using Variant Effect Predictor, with population frequencies from gnomAD and Bravo/TOPMed freeze 5 collections.^32^ All putative causal variants were manually inspected in IGV. Capillary sequencing for *MFRP* or *PRSS56* was performed using primer sequences available upon request. RNAseq data was generated on tissues from 21 healthy donor eyes from 21 individuals as previously described.^33^

### Statistical analysis

Data were analysed with R (v3.6.1), with continuous variables displayed as mean ± SD (for normally distributed variables) or IQR (for non-normal variables), and categorical variables displayed as numbers and percentages. Mean values for axial length and spherical equivalent were calculated per individual using data from both eyes. *P* values were calculated by one-way ANOVA with Tukey multiple pairwise-comparison testing for normally distributed continuous variables, by Kruskal-Wallis test for non-normal continuous variables, or by Fisher’s exact test for categorical variables. One-way between-group analysis of covariance was used to compare differences between groups adjusted by age.

## Results

We recruited a total of 40 individuals with nanophthalmos from 13 families. The mean age at recruitment was 47.5 ± 20.7 years, 57.5% were female, and 69.2% of families were of self-reported European ancestry (European (9), mixed European/Indigenous Australian (1), Lebanese (1), Vietnamese (1), not reported (1)) (Figure 1A). Exome sequencing was performed on one or more affected individuals from each family, with the exception of family NNO03, in which a causative variant was identified by capillary sequencing. When including two previously reported pedigrees^23,28^, a suspected genetic cause was identified in 9 of 13 kindreds (69.2%), in genes including *PRSS56* (4), *MFRP* (3), *TMEM98* (1), and *MYRF* (1) (Figure 1B, C, D, E). All variants were rare or absent in gnomAD, with CADD scores above 24, and recessive variants confirmed to be *in trans* by parental segregation testing (Table 1). Within tissues of dissected cadaveric human eyes^33^, expression of the dominant genes *MYRF* and *TMEM98* was more similar to one another that to the recessive genes *MFRP* and *PRSS56* (Figure 1F).

**Table 1.** Disease-associated gene variants in nine nanophthalmos probands. Genomic coordinates are based on the hg19 reference. AF, gnomAD r2.1.1 allele frequency; CADD, Phred-scaled CADD score. Variant consequences refer to the following transcript and protein accession IDs: *TMEM98* (ENST00000579849.1, ENSP00000463245.1), *PRSS56* (ENST00000617714.1, ENSP00000479745.1), *MFRP* (ENST00000555262.1, ENSP00000450509.1), *MYRF* (ENST00000278836.5, ENSP00000278836.4).

| Family | gene | chr | start | ref | alt | state | class | cDNA | protein | AF | CADD |
| --- | --- | --- | --- | --- | --- | --- | --- | --- | --- | --- | --- |
| NNO03 | *MFRP* | 11 | 119214525 | C | A | het | splice donor | c.1124+1G>T | . | 0.00006065 | 26.4 |
| NNO03 | *MFRP* | 11 | 119212289 | G | A | het | missense | c.1709C>T | p.Ala570Val | 0.000004098 | 24.4 |
| **NNO04** | *MFRP* | 11 | 119212574 | C | A | het | missense | c.1508G>T | p.Gly503Val | . | 25.6 |
| **NNO04** | *MFRP* | 11 | 119216248 | G | A | het | stop gained | c.523C>T | p.Gln175Ter | 0.00004022 | 36 |
| **NNO08** | *MFRP* | 11 | 119214525 | C | A | hom | splice donor | c.1124+1G>T | . | 0.00006065 | 26.4 |
| **NNO32** | *MYRF* | 11 | 61553331 | C | - | het | frameshift deletion | c.3361del | p.Arg1121GlyfsTer36 | . | . |
| **NNO02** | *PRSS56* | 2 | 233387913 | G | T | het | splice donor | c.849+1G>T | . | 0.00008335 | 24.5 |
| **NNO02** | *PRSS56* | 2 | 233388527 | - | C | het | frameshift insertion | c.1066dupC | p.Gln356ProfsTer152 | 0.0003668 | . |
| **NNO18** | *PRSS56* | 2 | 233388527 | - | C | hom | frameshift insertion | c.1066dupC | p.Gln356ProfsTer152 | 0.0003668 | . |
| **NNO22** | *PRSS56* | 2 | 233385406 | - | T | hom | splice donor | c.97+2dupT | . | . | . |
| **NNO27** | *PRSS56* | 2 | 233386763 | - | G | het | frameshift insertion | c.343dupG | p.Ala115GlyfsTer39 | 0.000009972 | . |
| **NNO27** | *PRSS56* | 2 | 233390091 | C | T | het | missense | c.1690C>T | p.Arg564Cys | 0.00001887 | 33 |
| **NNO01** | *TMEM98* | 17 | 31267907 | G | C | het | missense | c.577G>C | p.Ala193Pro | . | 29.5 |

**Figure 1:**
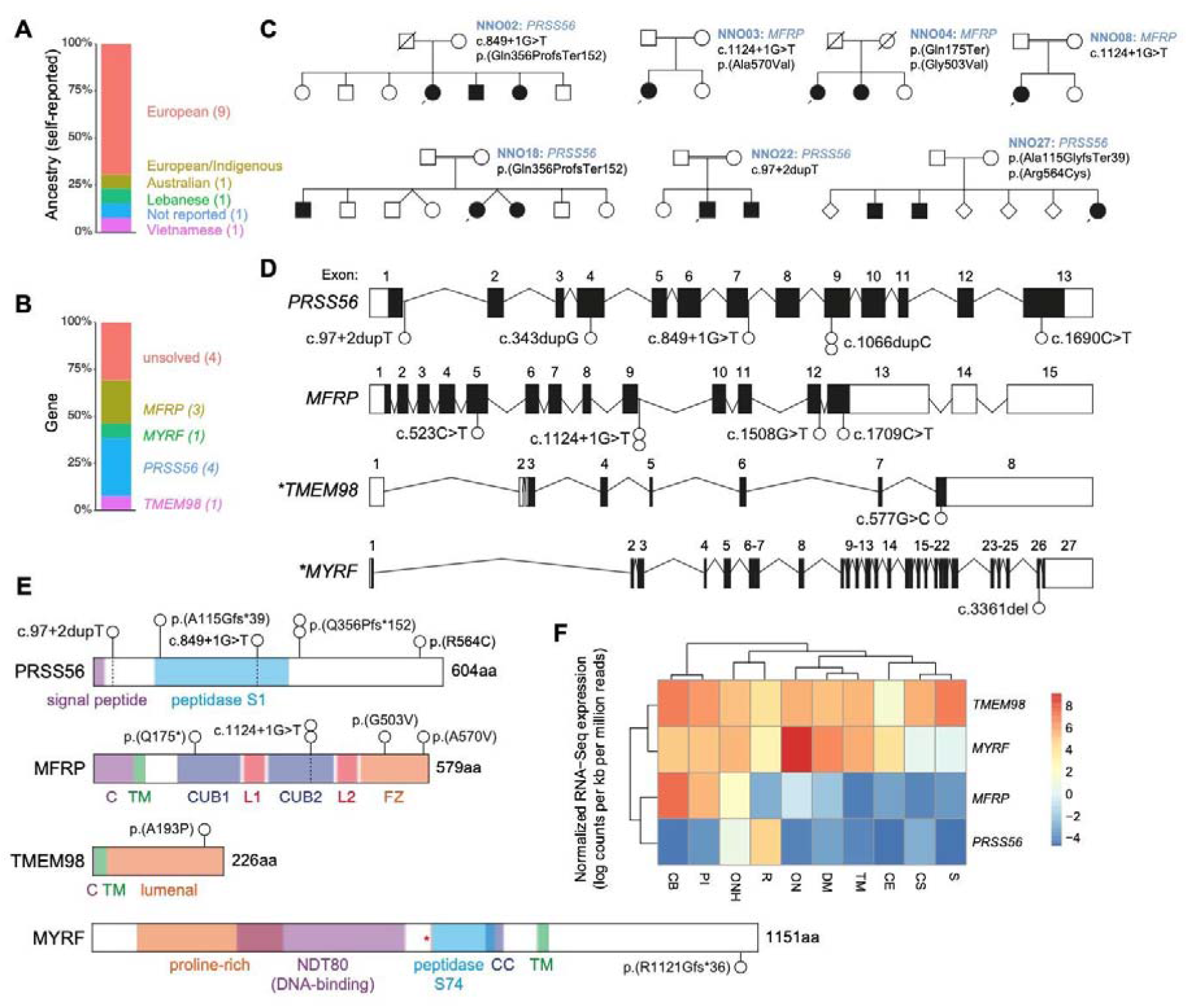
Genetic characterisation of a nanophthalmos cohort. (A) Self-reported ancestry of the 13 probands. (B) Proportion of the cohort assigned likely pathogenic variants, and genes in which they occur. (C) *PRSS56*- and *MFRP*-associated pedigrees. Square symbols, male; round symbols, female; diagonal line, deceased; black filled symbols, affected (nanophthalmos); unfilled symbols, unaffected; black arrow, proband; double horizontal line, consanguinity. Genotypes of affected individuals displayed alongside each family identifier. (D) Schematic of the *PRSS56, MFRP, TMEM98*, and *MYRF* loci, indicating the location of variants identified in the current study. Filled boxes indicate coding sequence, asterisks indicate genes with variants from previously reported pedigrees. (E) PRSS56, MFRP, TMEM98, and MYRF protein schematics, including domain structures and relative location of variants identified in the current study (white circles). Dashed vertical lines correspond to exon boundaries. C, cytoplasmic domain; TM, transmembrane domain; CUB, Complement C1r/C1s, Uegf, Bmp1 domain; L, LDL-receptor class A domain; FZ, frizzled domain; CC, coiled-coil domain. (F) Relative mean expression (expressed a log counts per kb per million mapped reads) of four known nanophthalmos genes (*TMEM98, MYRF, MFRP, PRSS56*) in dissected human adult cadaveric eye tissue. S, sclera; CS, corneal stroma; CE, corneal epithelium; TM, trabecular meshwork; DM, Descemet’s membrane; ON, optic nerve; ONH, optic nerve head; PI, peripheral iris; CB, ciliary body.

We identified four predicted pathogenic variants in *MFRP* across three probands, including one nonsense variant previously associated with nanophthalmos (p.(Gln175Ter))^11^, one splice site variant previously associated with retinitis pigmentosa (c.1124+1G>T)^34^, and two missense variants with no previous disease associations (p.(Ala570Val) and p.(Gly503Val)) (Figure 1D,E).

Of the four probands with variants in *PRSS56*, all were found to have rare compound heterozygous or homozygous variants (Figure 1D,E, Table 1). These variants include previously unreported essential splice (c.849+1G>T and c.97+2dupT), frameshift (p.(Ala115GlyfsTer39)), and missense variants (p.(Arg564Cys)) predicted to be pathogenic, as well as two instances of the previously reported c.1066dupC frameshift variant. The *PRSS56* c.1066dupC frameshift insertion (also reported as c.1059_1066insC, or p.(Gln356ProfsTer152)), had previously been reported in multiple Tunisian families with posterior microphthalmos (Figure 2A).^15,17,35^ A more recent study revealed that a c.1066dupC variant found in multiple Tunisian families was derived from a common ancestor around 1,850 years earlier.^35^ Another nine cases explained by the c.1066dupC variant have also been reported in a Saudi Arabian cohort of microphthalmia and posterior microphthalmos, accounting for 69% (9/13) of all cases with *PRSS56* variants in this report^8^, as well as in Egyptian and Lebanese patients.^36^ It is not clear whether this was also due to a founder effect, although cohort ancestry (Saudi Arabian, Egyptian, and Lebanese) suggests that these cases were unlikely to share the Tunisian founder haplotype.

**Figure 2:**
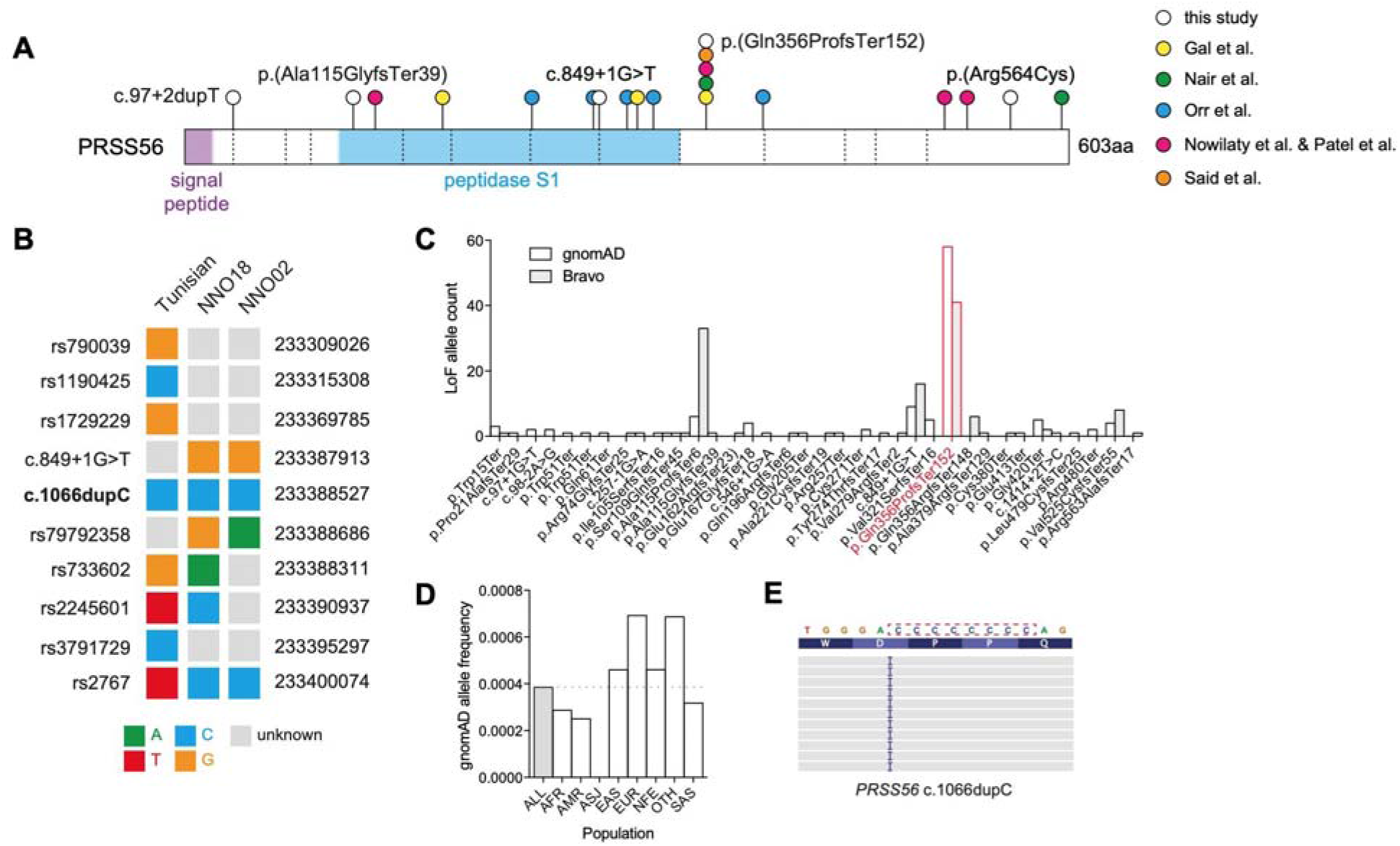
The origins of *PRSS56* c.1066dupC in a mutational hotspot. (A) Schematic of the PRSS56 protein, including domain structures and the relative location of variants identified in the current study (white circles), or in previous reports (coloured circles). Dashed vertical lines correspond to exon boundaries. (B) Comparative haplotype structure surrounding the *PRSS56* c.1066dupC variant in the previously reported Tunisian founder, and two unrelated probands reported here (NNO18, NNO02). (C) Frequency of *PRSS56* loss-of-function variants (nonsense, essential splice, frameshift) reported in the gnomAD r2.0.2 and Bravo (TOPMed Freeze5) collections. The c.1066dupC variant is highlighted in red. (D) Allele frequency of the c.1066dupC variant across multiple ancestries within gnomAD r2.0.2. (E) IGV representation of the left-shifted c.1066dupC variant in the context of the cytosine mononucleotide repeat.

To determine the origin of the c.1066dupC frameshift variant in our two Australian kindreds, we compared surrounding haplotypes in NNO18, NNO02, and the previously described Tunisian founder haplotype (Figure 2B).^35^ Three phased variants were genotyped in both the NNO18 proband and the Tunisian founder (rs733602, rs2245601, and rs2767), yet the genotypes in each were divergent, suggesting that the two alleles had arisen independently. Similarly, the NNO02 haplotype included the C allele of rs2767 (compared to T in the Tunisian founder), suggesting that the c.1066dupC variant in NNO02 was also unlikely to have come from the Tunisian founder. To compare the origins of the *PRSS56* haplotypes in NNO02 and NNO18, we made use of a closely linked variant (rs79792358) in *PRSS56* (149 bp downstream of c.1066dupC). This variant and the c.1066dupC insertion were both present in the NNO02 proband but absent from the NNO18 proband, suggesting that the NNO02 and NNO18 frameshift variants arose independently on different haplotypes (Figure 2B).

The c.1066dupC frameshift insertion was the most frequent loss-of-function *PRSS56* variant in the gnomAD collection of genomes and exomes (Figure 2C), representing over half (58/112) of all reported *PRSS56* loss-of-function variants. These 58 c.1066dupC variants were distributed across seven ancestral populations at allele frequencies between 2.5 x 10^−4^ and 6.9 x 10^−4^, consistent with the independent emergence of c.1066dupC variants in multiple populations (Figure 2D). *PRSS56* c.1066 was also noted to be a multiallelic site, with insertions and deletions reported at the same position (Figure 2C), both of which altered the length of an eight nucleotide cytosine mononucleotide repeat (Figure 2E).

Individuals with suspected pathogenic variants in *PRSS56, MFRP, TMEM98*, or *MYRF* had reduced ocular axial length (Figure 3A, Table 2), with the majority also highly hyperopic (Figure 3B, Table 2). The mean axial length was significantly shorter in groups with a genetic diagnosis compared to those without (*P*=7.22×10^−9^), with the *PRSS56* subset also having a shorter mean axial length than the dominant *TMEM98* (adjusted *P*=0.00003) and *MYRF* (adjusted *P*=0.00004) subsets (Figure 3A). All groups also had a significantly higher mean refractive error (*P*=0.0005), with *PRSS56* and *MFRP* groups more hyperopic than nanophthalmic individuals without a genetic diagnosis (Figure 3B, Table 2). Across the entire cohort and in those where data were available, 22.6% (7/31) had glaucoma, and 43.3% (13/30) had a pressure-lowering surgical intervention in at least one eye (laser peripheral iridotomy, selective laser trabeculoplasty, or trabeculectomy). Neither of these variables showed a significant difference between the genetic diagnosis groups (*P*=0.6665 and *P*=1.0000 respectively), even after adjusting for age at recruitment (Table 2).

**Table 2.**
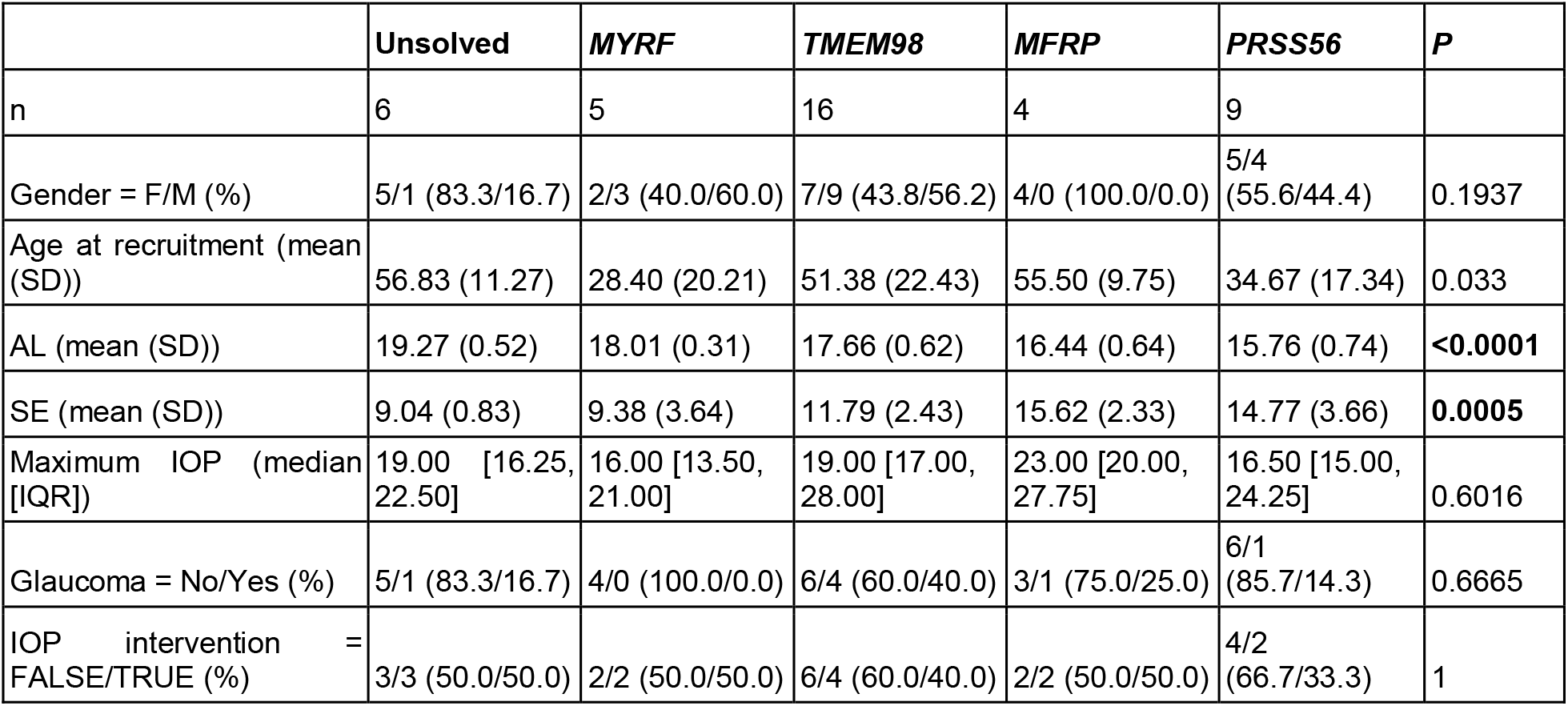
Summary of genotype-phenotype correlates. Continuous variables are displayed as mean ± SD (for normally distributed variables) or IQR (for non-normal variables), with categorical variables displayed as numbers and percentages. *P* values were calculated by one-way ANOVA for normally distributed variables, Kruskal-Wallis test for non-normal continuous variables, or by Fisher’s exact test for categorical variables. AL, axial length; AC, anterior chamber; SE, spherical equivalent; MD, mean deviation; SD, standard deviation; IQR, interquartile range.

**Figure 3:**
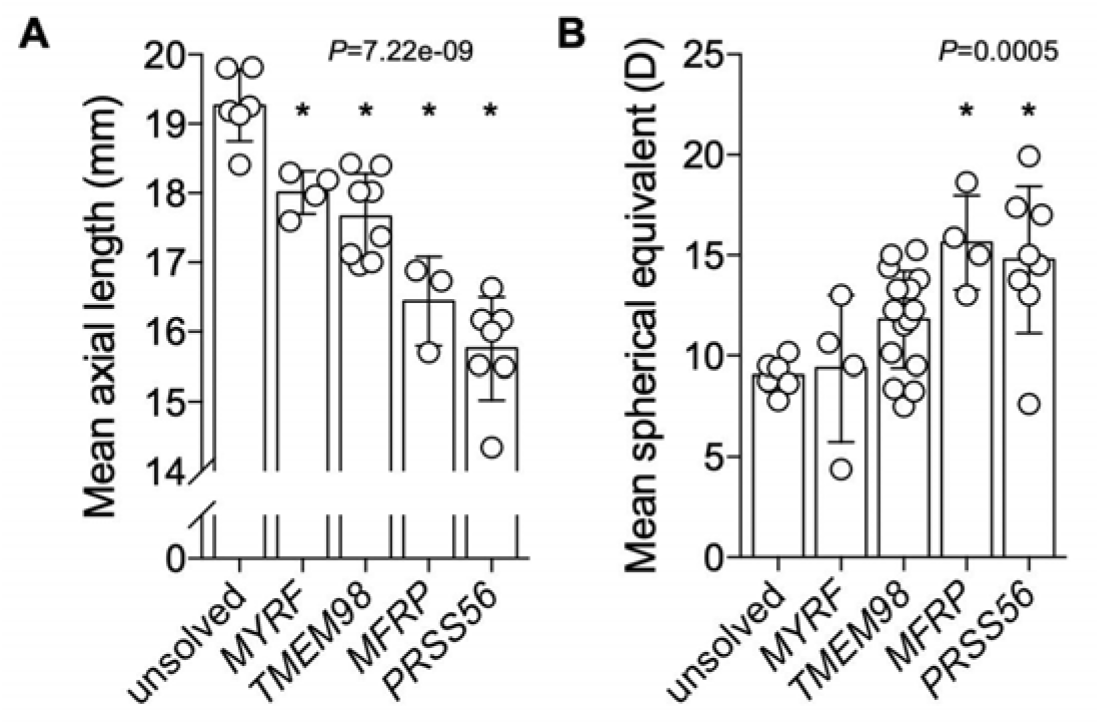
Genetic diagnosis is associated with shorter axial lengths and increased hyperopia. Ocular axial length (A) and spherical equivalent (B) of affected family members stratified by genetic diagnosis. Mean values were calculated per individual using data from both eyes. For both comparisons, group means were significantly different by one-way ANOVA (*P*<0.0003), with asterisks (*) indicating a significant difference from the unsolved cohort (Tukey multiple comparison testing, *P*<0.003).

## Discussion

Here we described an Australian nanophthalmos cohort of predominantly European ancestry in which 53.8% (7/13) of families were explained by variants in *MFRP* or *PRSS56*, or 69.2% (9/13) when including previously reported families with variants in *TMEM98* or *MYRF*.^23,28^ Mean axial length was shorter in all four groups of individuals with a genetic diagnosis compared to those without, with the recessive *PRSS56* and *MFRP* forms associated with the shortest axial length. Similarly, hyperopia was higher in affected individuals with a known genetic diagnosis compared to those without. This suggests that individuals with milder nanophthalmos phenotypes may be explained by variants in other genes, and emphasises the importance of prioritising genetic testing for *MFRP* and *PRSS56* in individuals with the most severe nanophthalmos phenotypes.

All individuals with an axial length below 18 mm in their smaller eye had a genetic diagnosis (17/17), with all but one under 17 mm associated with variants in either *PRSS56* or *MFRP* (10/11). Previous population studies have shown these thresholds to capture 0.137% (<18 mm) and 0.05% (<17 mm) of an adult British population^30^, and we propose that these be considered a threshold for identifying an underlying single gene cause.

*PRSS56* was the most commonly implicated gene in this cohort (4/13, 30.8%) and in other cohorts of posterior microphthalmos and nanophthalmos (27.3% - 61.5%).^8,26,36^ Including the four described here, at least 32 pedigrees have been described with *PRSS56*-associated posterior microphthalmos or nanophthalmos.^8,15–17,26,35,36^ Twenty of these pedigrees (62.5%) share the same c.1066dupC frameshift insertion variant, or 55.6% (15/27) if one considers the six Tunisian pedigrees with a shared haplotype as a single extended pedigree.^17,35^ Two of the families reported here harbour the c.1066dupC variant on at least one allele, which lay on haplotypes distinct from each other and from the previously reported Tunisian pedigree, suggesting the existence of a mutational hotspot in *PRSS56*. These variants arose in the context of an eight nucleotide cytosine mononucleotide repeat, which is incidentally a frequent occurrence in *MYRF*-associated nanophthalmos and high hyperopia^28^, and is likely secondary to slipped-strand mispairing during DNA replication.^37^

While loss-of-function variants such as *PRSS56*:c.1066dupC are associated with reduced axial length and hyperopia, variants that enhance the activity of PRSS56 might be expected to lead to increased axial length and myopia. The p.(Ala30Thr) variant of *PRSS56* is one of the few protein-coding variants to have emerged from genome-wide association studies of refractive error. Unlike the loss-of-function variants described here which associate with hyperopia, p.(Ala30Thr) is associated with myopia, suggesting that it may increase axial length through a gain-of-function mechanism.^20^ PRSS56 itself is a soluble serine protease, potentially sensitive to protease inhibitors or monoclonal antibodies, and therefore would be a candidate therapeutic target in the treatment of refractive error. It has already been shown that deletion of *Prss56* can reduce axial length and correct refractive error in an *Egr1* mutant mouse model of myopia^18^, raising the possibility that PRSS56 inhibitors might be applied to the treatment of myopia in humans.

In summary, we present the genetic landscape of nanophthalmos in an Australian cohort, revealing a considerable overlap between genes associated with rare and common refractive error phenotypes. The shortest axial length and most extreme hyperopic refractive error were associated with autosomal recessive forms of nanophthalmos (*PRSS56* and *MFRP*), with a less extreme phenotype in autosomal dominant forms (*TMEM98* and *MYRF*). Finally, we demonstrated that the most frequent single variant associated with nanophthalmos across all ancestries, *PRSS56* c.1066dupC, is likely to have arisen independently in at least four different populations, highlighting the importance of a single mutational hotspot in the global prevalence of nanophthalmos.

## Data Availability

Anonymized data can be made available upon request.

## Acknowledgments

Supported by grants from the National Health and Medical Research Council (NHMRC APP1107098), the Ophthalmic Research Institute of Australia, and Flinders University. KPB was funded by an NHMRC Career Development Award, and JEC by an NHMRC Practitioner Fellowship. DGM was supported in part by the National Human Genome Research Institute, the National Eye Institute, and the National Heart, Lung and Blood Institute grant UM1 HG008900, and by the National Human Genome Research Institute grant R01 HG009141.

